# Long-term individual and population functional outcomes in older adults with atrial fibrillation

**DOI:** 10.1101/2020.11.05.20223297

**Authors:** Anna L. Parks, Sun Y. Jeon, W. John Boscardin, Michael A. Steinman, Alexander K. Smith, Margaret C. Fang, Sachin J. Shah

## Abstract

**Background:** Older adults with atrial fibrillation (AF) have multiple risk factors for disablement. Long-term function and the contribution of strokes to disability has not been previously characterized.

**Methods:** We performed a longitudinal, observational study in the nationally representative Health and Retirement Study (1992-2014). We included participants ≥65 years with Medicare claims who met incident AF diagnosis claims criteria. We examined the association of incident stroke with three functional outcomes: independence with activities of daily living (ADL) and instrumental activities of daily living (IADL) and community-dwelling. We fit separate logistic regression models with repeated measures adjusting for comorbidities and demographics to estimate the effect of stroke on function. We estimate the contribution of strokes to the overall population burden of functional impairment using the method of recycled predictions.

**Results:** Among 3530 participants (median age 79 years, 53% women, median CHA2DS2-VASc 5), 262 had a stroke over 17,396 person-years. Independent of stroke and accounting for population comorbidities, annually, ADL dependence increased by 4.4%, IADL dependence increased by 3.9%, and nursing home residence increased by 1.2% (p<0.05 for all). Accounting for comorbidities, of those who experienced a stroke, 31.9% developed new ADL dependence, 26.5% developed new IADL dependence, and 8.6% newly moved to a nursing home (p<0.05 for all). Considering all causes of function loss, 1.7% of ADL disability-years, 1.2% of IADL disability-years, and 7.3% of nursing home years could be attributed to stroke over 7.4years.

**Conclusion:** Older adults lose substantial function over time following AF diagnosis, independent of stroke. Stroke was associated with a significant decline in function and an increase in the likelihood of nursing home move, but stroke did not accelerate subsequent disability accrual. Because of the high background rate of functional loss, stroke was not the dominant determinant of population-level disability in older adults with AF.

**Impact statement:** We certify that this work is novel. Little is known about long-term function (ADL, IADL, community-dwelling) among older adults with AF and the association with stroke. This nationally representative study finds a high rate of function loss independent of stroke, and among those who suffer a stroke, a dramatic and immediate decline in function. Because of the high rate of function loss independent of stroke and the relatively low rate of stroke, on a population level, stroke is not the dominant determinant of disability in older adults with AF.

## Introduction

Atrial fibrillation (AF) is a significant health burden for older adults, affecting 1 in 25 of those over age 60 and 1 in 10 over age 80.^1^ Stroke is the most dreaded consequence of AF for older adults, who often equate stroke with abrupt and persistent loss of independence.

While prior studies have clearly demonstrated that AF-related strokes result in increased short-term disability, long-term functional outcomes for AF patients generally, and specifically before and after stroke, remain uncharted.^2^ While many strokes result in sudden and life-altering disability, disability can also result from an accumulation of impairments independent of stroke. Particularly in older adults, AF frequently coexists with other medical conditions that increase the risk of disability.^3,4^ Also, while often overlooked, geriatric syndromes, such as falls and cognitive impairment, are also important contributors to loss of independence in older AF patients.^5,6^ In large and nationally representative cohorts of older adults with AF, frailty, cognitive impairment, and functional impairments are common.^7,8^ In the absence of longitudinal data in older adults with AF, the contribution of stroke and factors besides stroke to long-term disablement remains unclear.

Stroke is perceived as a dominant pathway to disability in patients with AF, yet information assessing its actual population health burden is limited. Compared to the prevalence of other risk factors for disability, stroke remains an uncommon occurrence even among AF patients. Thus, many older adults with AF may be predisposed to disablement even in the absence of stroke, but this has not been previously quantified.

Understanding the longitudinal course of independence of older patients with AF, and whether and to what degree stroke affects their long-term trajectory, is vital to inform treatment decision-making, advance care planning, and public health interventions. To address these gaps, we used a nationally representative cohort of older adults with incident AF to determine long-term functional outcomes and the relative contribution of strokes.

## Methods

### Study design and participants

We performed a longitudinal, observational study to examine the association between incident stroke and functional status in a cohort of older adults with AF. We used data from 1992-2014 from the Health and Retirement Study (HRS), a longitudinal, nationally representative survey of more than 37,500 Americans age 50 and older, which amounts to 350,000 person-years of observation.^9^ Subjects are interviewed every two years by phone, in person or via internet surveys. The goal of the HRS is to measure changes in health, wealth, social structure, and function as participants retire, with questions covering four aging-related topics: economic security, mental and physical health and function, work and retirement, and social connections. Proxy interviewees, typically family members, are used for those who cannot participate because of physical or cognitive impairment. The HRS is linked to Medicare insurance claims for participants who consent.^10^ The HRS sampling design yields a nationally representative sample of community-dwelling adults and nursing home residents in the US.^11^

We identified a cohort of HRS subjects aged 65 and older with Medicare claims linkage and continuous Medicare Part A and B enrollment (cohort flow diagram **Appendix 1**). We included participants if their claims had one inpatient or two outpatient claims for AF in the first or second position (427.31 from the *International Classification of Diseases Ninth revision, Clinical Modification*). Cases were defined as incident AF if they had 12 months of continuous enrollment in Medicare Part A and B with no prior AF claims diagnosis.^12^ To ensure accurate information about participants’ baseline characteristics, we excluded those whose first interview occurred more than 2.5 years before their AF diagnosis; a timeframe of 2.5 years was selected because of the variable biennial timing between HRS interviews.^13^ To obtain longitudinal outcome and exposure information, we excluded participants who had no interviews after a diagnosis of AF. Participants were censored at the time of disenrollment from Medicare part A or B, HRS drop out, or death, whichever came first.

### Measures: Demographics and comorbidities

We used HRS interview data to characterize participants’ age, gender, education level, race, ethnicity, education, marital status, and whether they lived alone. We report information on comorbid conditions that affect stroke risk, including congestive heart failure, hypertension, prior stroke, diabetes mellitus, prior myocardial infarction or angina, or cancer (excluding skin cancer). We characterized patients as having a specific clinical comorbidity if (1) they report a physician had ever told them they had a specific condition or (2) they met Medicare claims criteria (definitions found in **Appendix 2**). Prior studies have examined the validity of self-reported cardiovascular comorbidities, such as those used in the CHA2DS2-VASc score, finding self-reported diagnoses to reflect medical charts and population-level estimates accurately.^14–16^ Clinical characteristics were used to compute a CHA2DS2-VASc (Congestive heart failure, Hypertension, Age, Diabetes mellitus, Stroke, Vascular disease, and Sex) score to estimate the risk of ischemic stroke.^17^ We report participants’ baseline demographic and comorbidity information from the most recent interview before their AF diagnosis.

### Measures: Stroke

We used a claims-based definition of ischemic (ICD-9 codes: 434.x and 436.x; ICD10: I63.x) and hemorrhagic stroke (ICD9: 430.x and 431.x; ICD10: I61.x).^18^ To classify incident stroke events, we identified subjects with an inpatient admission with a primary diagnosis code for ischemic or hemorrhagic stroke. We examined the first stroke after AF diagnosis for each participant in the analysis.

### Outcomes: Function

We measured the effect of strokes on three functional outcomes: activities of daily living (ADL) impairment, instrumental activities of daily living (IADL) impairment, and community dwelling. We defined ADL dependence as a respondent stating that they required help with any of 6 ADLs (walking, dressing, bathing, eating, transferring to or from bed, and toileting). We defined IADL dependence as a respondent stating that they required assistance with any of 5 IADLs (preparing a hot meal, shopping for groceries, making telephone calls, taking medicines, and managing money). We defined nursing home move as spending ≥90 nights in a nursing home since the last interview. We defined the outcome of community-dwelling as no nursing home move since the last interview. All three functional outcomes were measured at each interview.

### Analysis

We measured the prevalence of cohort demographics and comorbidities at the time of AF diagnosis. To determine the association between stroke and each functional outcome, we fit separate random-effects logistic regression models with repeated measures. We modeled the effect of stroke on function by including an interaction term between stroke and time since AF diagnosis (modeling equation in **Appendix 3**). In each model, we adjusted for covariates, defined a priori, that contribute to stroke and disability: age, sex, congestive heart failure, hypertension, prior stroke, diabetes mellitus, prior myocardial infarction or angina, or history of cancer (excluding skin cancer). We determined the log-odds for each outcome: ADL dependence, IADL dependence, and nursing home residence. To visually illustrate the effect of stroke on the probability of ADL independence, IADL independence, and nursing home residence, we present the predicted population rates and the average marginal effects.^19^ We did not use significance testing to determine which confounders to include in the regression models, consistent with epidemiologic best practices.^19^ If, during an otherwise completed interview, ADL, IADL, nursing home status were not obtained, the participant was not included in the analysis for that outcome in that time period (<4% of interviews). A substantial proportion of missing interviews were exit interviews with next-of-kin (1.5% of all interviews). When an exit interview was completed, next-of-kin reported ADL and IADL dependence >90% of the time. As such, when exit interviews were missing, we imputed ADL and IADL status as dependent.

To estimate the contribution of strokes to the overall population burden of functional impairment in patients with AF, we used the method of recycled predictions.^20,21^ First, using the regression model results (**Appendix 4)** and the observed stroke rate, we estimated the burden of dependence for the population that had a stroke and the population that did not have a stroke projected over 7.4 (measured in disability-years). We chose 7.4 years because it was the 75^th^ percentile of follow-up time. Next, we used the same parameters to estimate the disability-years for the same population assuming no strokes had occurred. The difference between the two measures of disability-years represents the disability-years attributable to stroke (detailed methods and visual description presented in **Appendix 5**).

We performed all analyses using SAS 9.4 (Cary, NC) and STATA (Version 16.1, College Station, TX). We report all results with 95% confidence intervals. STROBE statement checklist can be found in **Appendix 6**.

## Results

### Cohort characteristics

We included 3,530 participants in our analysis with an average of 4.9 years of follow-up time, amounting to 17,396 person-years of observation. At the time of AF diagnosis, the median age was 79 years, 53% were women, 85% identified as white, 9.0% as Black, and 10.0% were proxy interviews (**Table**). The most common comorbid condition among participants was hypertension (79%), followed by prior myocardial infarction or angina (58%), congestive heart failure (36%), diabetes (31%), prior stroke (23%), and cancer (22%). The median CHA2DS2-VASc score in the cohort was 5 (IQR 4, 7). During the follow-up period after AF diagnosis, 262 participants, or 7.4%, had a stroke (1.5 strokes per 100 person-years). The median time from AF diagnosis to stroke was 2.8 years (IQR 0.8, 6.0).

**Table.** Baseline characteristics of the cohort of older adults with atrial fibrillation. QR - interquartile range; CHA_2_DS_2_-VASc score - congestive heart failure/hypertension/age/diabetes/stroke/vascular disease * none with CHA2DS2-VASc score of 0 Characteristics from baseline interview prior to AF diagnosis

|  | Prevalence (95% CI)<br>n=3530 |
| --- | --- |
| **Sociodemographics** |  |
| Age, median (IQR) | 79 (72, 85) |
| Women | 53% (52, 55) |
| Married or Partnered | 53% (52, 55) |
| Lives alone | 31% (30, 33) |
| Greater than high school education | 68% (67, 70) |
| Race |  |
| White | 85% (84, 86) |
| Black | 9% (8, 10) |
| Other | 2% (1, 2) |
| Hispanic ethnicity | 5% (4, 5) |
| Proxy interview | 10% (9, 12) |
| **Medical comorbidities** |  |
| Congestive heart failure (%) | 36% (34, 38) |
| Hypertension (%) | 79% (77, 80) |
| Diabetes mellitus (%) | 31% (30, 33) |
| Prior stroke (%) | 23% (22, 25) |
| History of vascular disease (%) | 58% (56, 59) |
| Cancer (%) | 22% (20,23) |
| CHA<sub>2</sub>DS<sub>2</sub>-VASc, median (IQR)* | 5 (4, 7) |
| 1 | 2% (1,2) |
| 2 | 5%(5,6) |
| 3 | 11% (10,12) |
| 4 | 18% (16,19) |
| 5 | 21% (19,22) |
| 6 | 18% (17,20) |
| 7 | 13% (12,14) |
| 8 | 9%(8,10) |
| 9 | 3% (3,4) |
| **Function at baseline** |  |
| ADL disability (%) | 16% (15, 17) |
| IADL disability (%) | 22% (20, 23) |
| Nursing home residence (%) | 3% (2, 3) |

### Longitudinal functional outcomes

Figure 1 graphically represents three key findings for each of the three functional outcomes— the baseline rate of lost function independent of stroke, the immediate loss of function following stroke, and change in the baseline rate of lost function following stroke. We present the full model results, including unadjusted results in **Appendix 4**.

**Figure 1:**
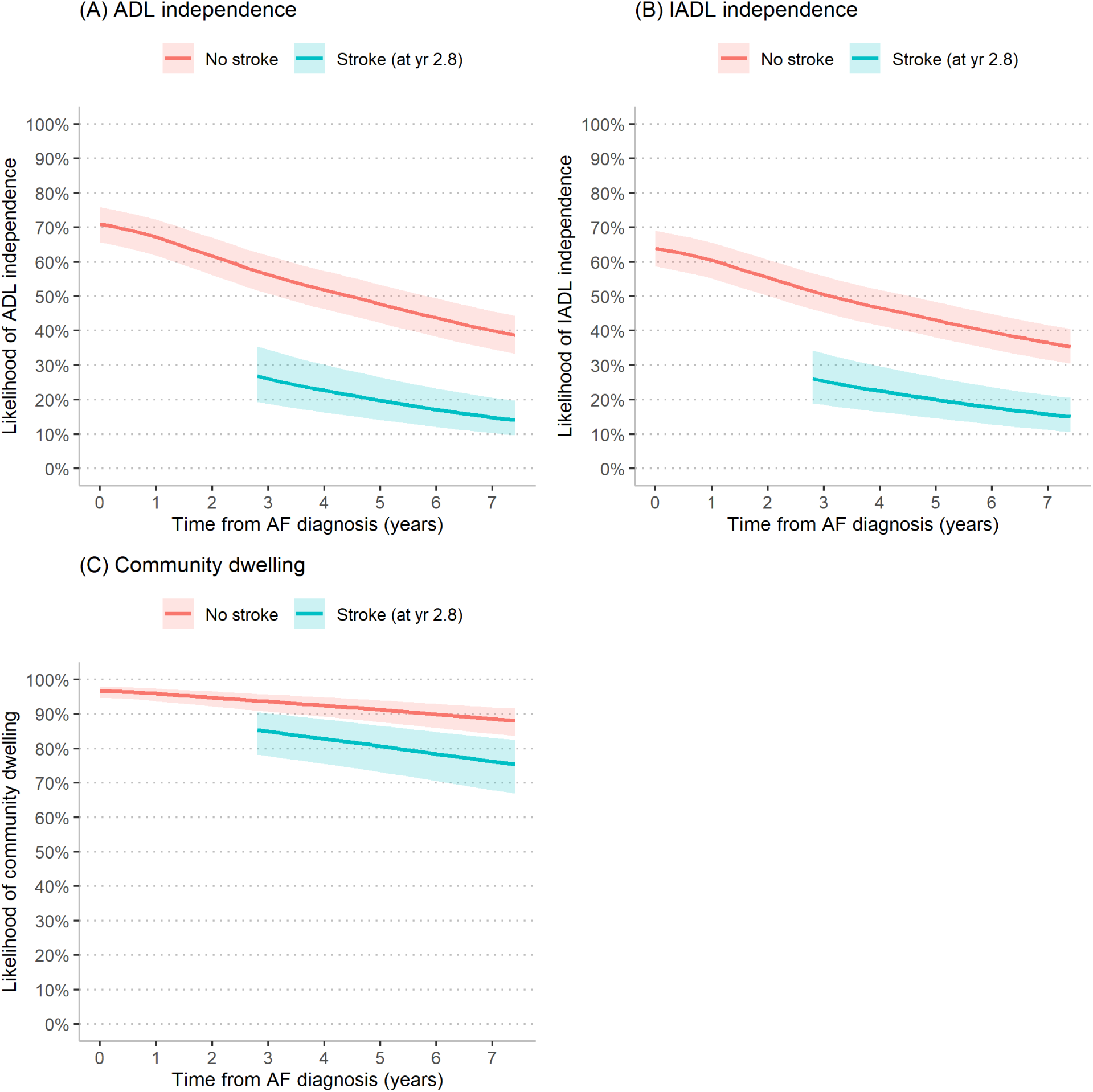
Longitudinal likelihood of ADL independence, IADL independence, and community dwelling and association with stroke. ADL – activity of daily living; IADL – instrumental activity of daily living The slope represents the average marginal effect of a one-year change on independence, adjusting for demographics and comorbidities. The blue line represents the predicted likelihood of the outcome without stroke and the red line represents the predicted likelihood of the outcome following stroke at 2.8 years (the median time to stroke in the cohort). For the population who goes on to have a stroke, the blue line from 0 to 2.8 years represents the pre-stroke trajectory and the red line describes the post-stroke trajectory. Functional trajectory is displayed through 7.4 years, the 75^th^ percentile of follow up time. The analysis is based on 3530 participants; we excluded 26 person waves (0.3%) with missing ADL outcome data, 213 person-waves (2.1%) with missing IADL outcome data, and 368 person-waves (3.5%) with missing nursing home outcome data.

Independent of stroke and accounting for comorbidities, the absolute likelihood of ADL independence decreased by 4.4% per year (average marginal effect [AME], 95% CI 4.0 to 4.8% per year) (**Figure 1A**). Assuming a stroke at 2.8 years, the median time to stroke in this cohort, stroke was associated with a decrease in the predicted likelihood of independence from 58% (pre-stroke) to 27% (post-stroke) (AME −32%, 95% CI -25 to -39%). Stroke, however, was not associated with a change in the baseline rate of decline in ADL independence; that is, the relative rate of ADL decline post-stroke did not differ when compared to those without stroke.

Similar to ADL independence, independent of stroke and accounting for comorbidities, the absolute likelihood of IADL independence decreased by 3.9% per year (AME, 95% CI 3.5 to 4.3% per year) (**Figure 1B**). Assuming a stroke at 2.8 years, stroke was associated with a decrease in the predicted likelihood of IADL independence from 52% to 26% (AME -27%, 95% CI -20 to -33%). Stroke was not associated with a change in the baseline annual rate of decline in IADL independence.

Finally, for community-dwelling, independent of stroke and accounting for comorbidities, the absolute likelihood of community-dwelling decreased by 1.2% per year (AME, 95% CI 1.0 to 1.4% per year) (Figure 1C). Assuming a stroke at 2.8 years, stroke was associated with a decrease in the predicted likelihood of community-dwelling from 94% to 85% (AME -8.6%; 95% CI -3.7 to -13.5%). Stroke was not associated with a change in the baseline annual rate of decline in community-dwelling.

### Population-level outcomes

Projected over 7.4 years, stroke accounted for a modest proportion of dependent-years among older adults with AF (**Figure 2**). Stroke accounted for 1.7% (95% CI 1.3 to 2.1%) of the population burden of ADL disability (for 100 person-years, 46.4 dependent-years given observed stroke rate vs. 45.6 disability-years assuming no strokes; attributable burden 0.8 disability-years/100 person-years or 1.7%). For IADL impairment, 1.2% (95% CI 0.9 to 1.6%) of the population burden of IADL disability could be attributed to stroke (for 100 person years, 51.8 dependent-years given observed stroke rate vs. 51.1 dependent-years assuming no strokes; attributable burden 0.6 dependent-years/100 person-years or 1.2%). For loss of community-dwelling, 7.3% (95% CI 4.0 to 10.5%) of the population burden of nursing home years could be attributed to stroke (for 100 person-years, 7.7 nursing home-years given observed stroke rate vs. 7.2 nursing home-years assuming no strokes; attributable burden 0.6 nursing home-years/100 person-years or 7.3%).

**Figure 2:**
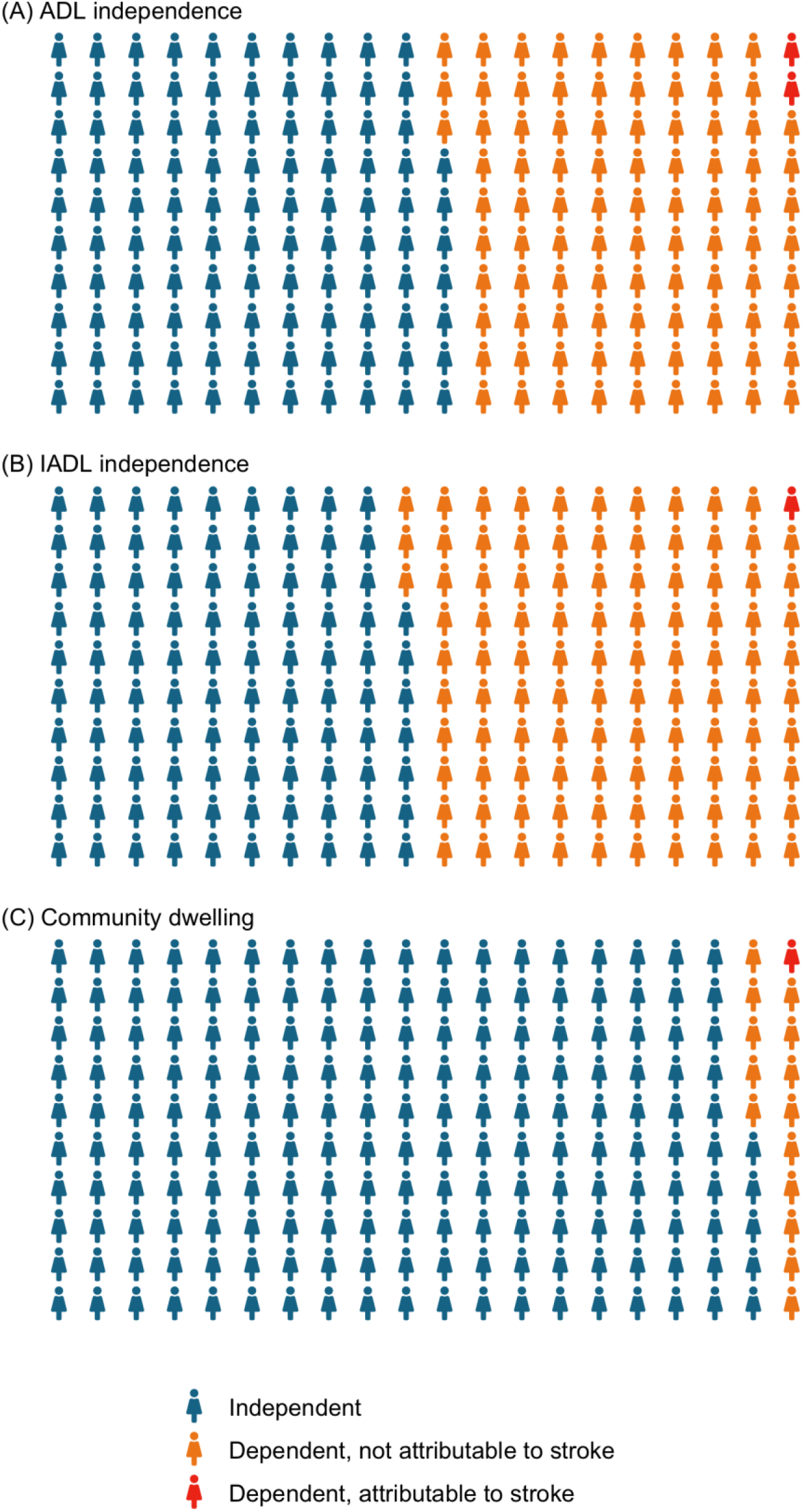
Contribution of strokes to population ADL independence, IADL independence, and community-dwelling over 100 person-years. ADL – activity of daily living; IADL – instrumental activity of daily living Each icon represents 0.5 person-years. Each panel contains 200 icons summing to 100 person-years. Values are all rounded to the nearest 0.5. Exact values are presented in the text of the results. To estimate the population burden of dependence attributable to strokes, we used the regression model results to calculate the likelihood of independence for each person based on their comorbidities and accounting for their survey sampling weight over seven years. Next, we used the same parameters to estimate the dependent-years for the same population assuming no strokes had occurred. The difference between the two measures of dependent-years represents the dependent-years attributable to stroke over 100 person-years (red icons, each representing 0.5 person-years). The blue icons represent independent 0.5 person-years. The yellow icons represent 0.5 dependent-years, not attributable to stroke.

## Discussion

In this nationally representative cohort, we determined that older adults with AF experience a significant loss of function over time. First, following AF diagnosis and independent of stroke, we found a high rate of loss of function and nursing home moves—annual increases of 4.4% in ADL dependence, 3.9% increase in IADL dependence, and 1.2% annual increase in nursing home residence. Second, stroke was associated with an immediate and substantial decline in function and an increase in the likelihood of nursing home move. In the years following a stroke, stroke survivors and those who did not experience a stroke accrued disability at similar rates; that is, the loss of independence did not accelerate following stroke.

Contrary to prevailing wisdom, we found stroke was not the dominant determinant of disability in older adults with AF on a population level. While a stroke results in striking loss of function for the 1 in 13 that suffer a stroke, we found the high background rate of function loss independent of stroke is the predominant pathway for population disability. In this AF cohort, projected over seven years, stroke accounted for 1.7% of ADL disability-years, 1.2% of IADL disability-years, and 7.3% of total time in nursing homes. Although AF-related stroke causes substantial, sudden loss of function for individuals and represents a substantial public health burden, most AF patients become disabled through other means.

These findings add to limited literature describing long-term functional outcomes among older adults with AF. Two recent studies have compared the risk of disability among AF patients versus those without AF. Marzona et al. showed that incident AF was associated with increased loss of ADL independence, as well as rates of admission to long-term care facilities, compared to those without AF.^22^ These associations were independent of stroke. However, because this study drew from patients enrolled in randomized controlled trials, with mean age a decade younger than in the current study, its results may be less generalizable to the older AF population. In a cohort of adults over age 65, Wallace et al. demonstrated that, even after adjusting for incident stroke or heart failure, incident AF was associated with 50% shorter disability-free survival and 24% higher risk of ADL disability than those without AF.^23^

The results of this study make clear that older adults with AF acquire disabilities through multiple mechanisms. Thus, mitigating disability may require distinct interventions. To ameliorate stroke-related disability, optimizing anticoagulant use and left atrial closure devices continues to be a primary concern.^24,25^ While this study demonstrates the importance of stroke-independent disability, the causes of stroke-independent disability are not well defined. Some have postulated a direct link between the dysrhythmia itself and function independent of stroke. For example, alterations in cardiac structure resulting from AF impair cardiac output could reduce patients’ capacity for both cognitive and physical activities.^26^ In addition to clinically diagnosed stroke, AF has also been associated with subclinical ischemic events and resultant disability.^27,28^ In this scenario, strategies could target preventing the development of atrial fibrillation altogether or preventing its cardiac remodeling effects. Rather than a causal connection between AF and disability, AF may instead be a marker for multimorbidity, frailty, and other geriatric syndromes that predispose to impairment. In this model, novel geriatric care models that address functional impairment using multifaceted interventions may hold promise.^29^ In either circumstance, when counseling patients and families regarding a diagnosis of AF, our results should prompt a discussion of the risk of functional dependence in the years following diagnosis of AF that is only partially mitigated by stroke prevention.

Our study has several limitations that should be considered when interpreting the results. We relied on self-report for functional status measures, which are subject to recall bias, as subjects may be reluctant to acknowledge these limitations or may not recall them due to cognitive deficits. We excluded participants who lacked follow-up data, which could have preferentially excluded those with more significant functional impairments who had difficulty completing follow-up interviews. However, both limitations would likely result in underestimating the true burden of disability and bias our results toward the null. The HRS assesses functional status every two years, but functional deficits can develop rapidly, as such important changes could be missed with this interval. Also, several studies have shown that older adults can have periods of decline and recovery, but our data did not allow for this degree of granularity.^30^ Nevertheless, although older adults often recover from initial disability, an episode of disability is a strong predictor of chronic impairment, so the overarching trajectory of function illustrated in our cohort remains informative.^31^ Participants with the most severe or deadly strokes may be differentially censored. To mitigate this bias, we assumed that all participants who died and were missing outcome data were disabled before death. In a sensitivity analysis, we examined if there was a differential loss to follow-up following a stroke. We did not find a significant association between stroke and differential loss to follow-up (**Appendix 7**); this reduces the likelihood but does not eliminate this potential bias. Finally, Medicare Part D prescription data are only available from 2006 onward. Stroke severity and short-term disability are both reduced by receipt of anticoagulant or antithrombotic medications, so this represents an important unmeasured confounder in our results that should be evaluated in future studies.^32^

In conclusion, we found that long-term functional decline was common in older adults with AF and mostly occurred in the absence of stroke. After seven years, one-third of AF patients had ADL disability, one-half IADL disability, and one-tenth lived in a nursing home. Considering the public health implications of AF in older adults, our results demonstrate that although stroke represents a significant cause of functional decline, most disability results from other causes. These results challenge the traditional thinking that the long-term function of AF patients depends principally on stroke prevention. Taken together, our findings emphasize the need to supplement stroke prevention along with multifaceted interventions to prevent dependence.

## Data Availability

Data sharing: Researchers can apply to use the Health and Retirement Study (hrs.isr.umich.edu/) for access to the data use in this study. Code used to generate the cohort and perform the analyses can be found on github (https://github.com/sachinjshah).

## Contributors

ALP, SYJ, WJB, and SJS, were responsible for the study concept and design. ALP, MAS, AKS, and SJS obtained funding and supervised the study. All authors were involved in the acquisition, analysis, or interpretation of the data. SJY performed the statistical analyses. ALP and SJS drafted the manuscript, and all authors critically revised it for important intellectual content. The corresponding author attests that all listed authors meet authorship criteria and that no others meeting the criteria have been omitted. ALP, SYJ, and SJS had full access to all the data in the study and are the guarantors.

## Funding

This study was supported by the National Center for Advancing Translational Sciences (KL2TR001870), the National Institute on Aging (P30AG044281, K24AG068312, K24AG049057 & T32-AG000212), and the National Heart Lung and Blood Institute (K24HL141354).

## Sponsor’s Role

The funders had no role in study design, data collection and analysis, decision to publish, or preparation of the manuscript.

## Competing interests

All authors have completed the ICMJE uniform disclosure form at www.icmje.org/coi_disclosure.pdf (available on request from the corresponding author). MCF reports grants from NIH/NHLBI, during the conduct of the study. ALP, MAS, AKS report grants from NIH/NIA, during the conduct of the study. SJS reports grants from NIH/NCATS, during the conduct of the study. No financial relationships with any organizations that might have an interest in the submitted work in the previous three years; no other relationships or activities that could appear to have influenced the submitted work.

## Ethical approval

The Human Research Protection Program Institutional Review Board at the University of California, San Francisco, approved this study (IRB# 16-19185).

## Data sharing

Researchers can apply to use the Health and Retirement Study (hrs.isr.umich.edu/) for access to the data use in this study. Code used to generate the cohort and perform the analyses can be found on github (https://github.com/sachinjshah).

## Appendix

## Appendix 1: Cohort flow diagram

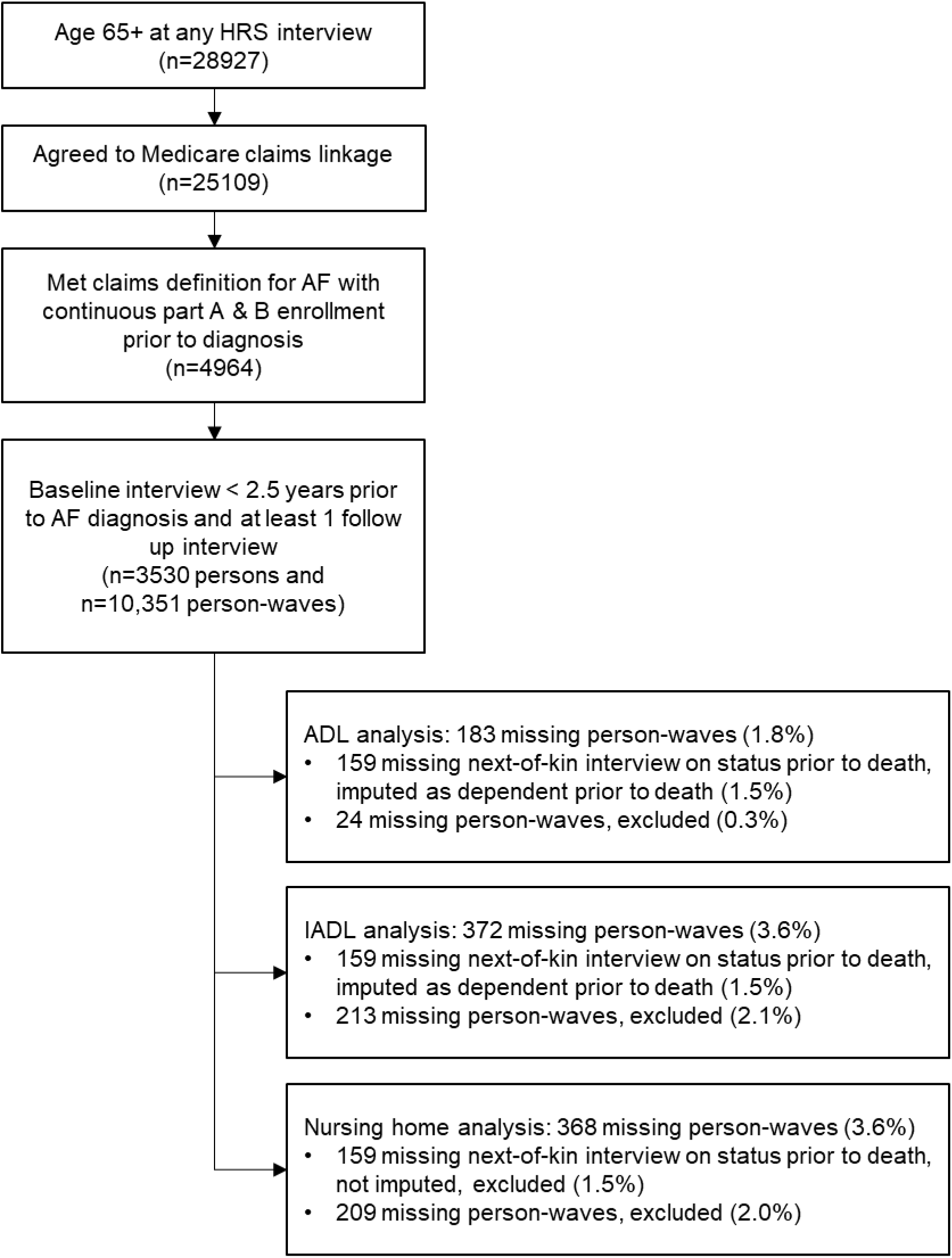

## Appendix 2: Medicare claims comorbidity definitions

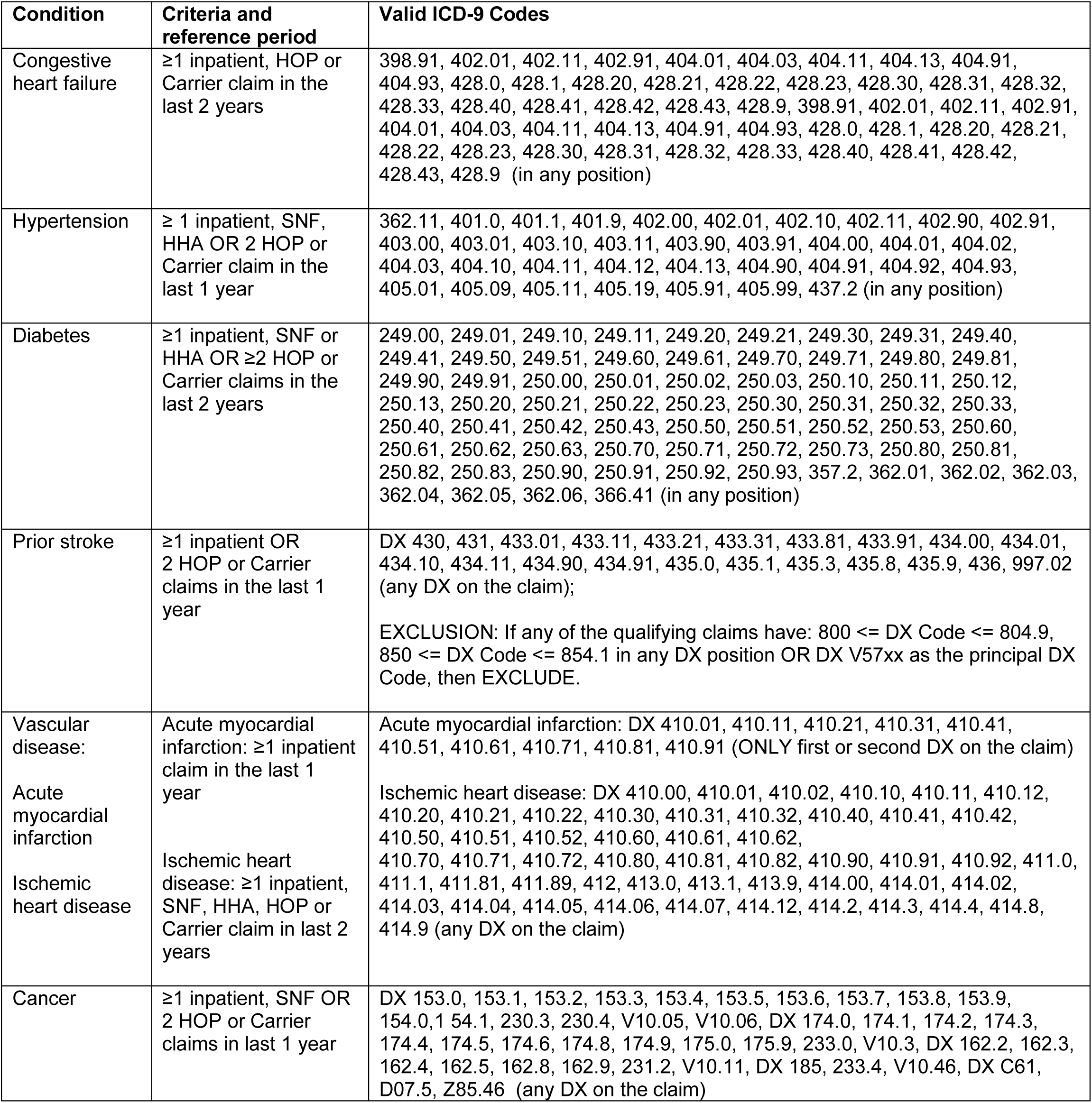

## Appendix 3: Modeling equation

We fit the following model random effects model using a binary distribution and logit link to estimate the association of stroke and function (ADL, IADL, and independent living).

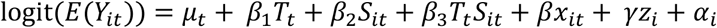

Where:

*Y*_*it*_= disability at time *t* for individual *i*

*T* = time since AF diagnosis

*S* = an indicator representing incident stroke and all time periods following an incident stroke

*x* = time-varying covariates (congestive heart failure, hypertension, diabetes, vascular disease, cancer, marital status, living alone, proxy respondent)

*Z* = time-invariant covariates (age at AF diagnosis, sex, history of stroke before AF diagnosis, race, ethnicity, educational attainment)

*α* _*i*_= unobserved individual specific effect

From this model, we determined the baseline relative risk of the outcome *β*_*1*_, the immediate change associated with stroke *β*_*2*_, and the change in baseline relative risk following stroke *β*_*3*_ These changes are graphically depicted below.

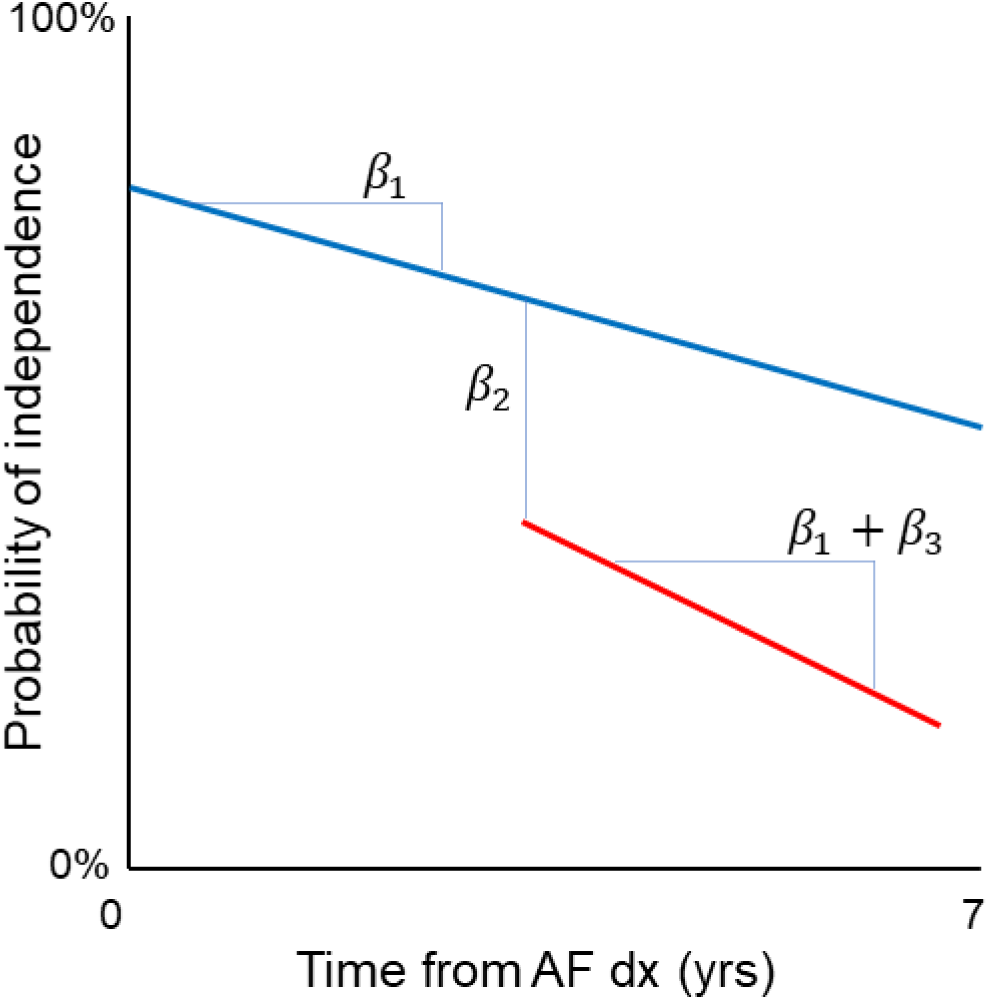

## Appendix 4: Manuscript Figure 1 results as table

Odds ratios and average marginal effects of stroke on ADL, IADL, nursing home residence adjusted for stroke risk

OR – odds ratio, AME – average marginal effect Unadjusted model does not include any time-varying or time invariant covariates. Adjusted model as noted in Appendix 4.

| Outcome | Parameter | Unadjusted OR<br>(95% CI) | Adjusted OR<br>(95% CI) | AME (95% CI) |
| --- | --- | --- | --- | --- |
| ADL<br>independence | Pre-stroke rate of incident disability $\beta_1$ | 0.67<br>(0.64-0.69) | 0.92<br>(0.89-0.95) | -4.4%/yr<br>(-4.0 to -4.8%/yr) |
| | Immediate change in disability following stroke $\beta_2$ | 0.02<br>(0.01-0.05) | 0.10<br>(0.05-0.21) | -31.9%<br>(-25.1 to -38.7%) |
| | Change in rate of incident disability post-stroke, relative to pre-stroke rate $\beta_3$ | 1.12<br>(1.01-1.25) | 1.02<br>(0.93-1.12) | -- |
| IADL<br>independence | Pre-stroke rate of incident disability $\beta_1$ | 0.74<br>(0.71-0.76) | 0.95<br>(0.92-0.97) | -3.9%<br>(-3.5 to -4.3%) |
| | Immediate change in disability following stroke $\beta_2$ | 0.04<br>(0.02-0.10) | 0.16<br>(0.08-0.32) | -26.5%<br>(-19.8 to -33.3%) |
| | Change in rate of incident disability post-stroke, relative to pre-stroke rate $\beta_3$ | 1.09<br>(0.99-1.20) | 1.02<br>(0.94-1.10) | -- |
| Continuous<br>independent<br>living | Pre-stroke rate of incident disability $\beta_1$ | 0.74<br>(0.71-0.78) | 0.96<br>(0.92-1.00) | -1.2%<br>(-1.0 to -1.4%) |
| | Immediate change in disability following stroke $\beta_2$ | 0.07<br>(0.03-0.18) | 0.24<br>(0.11-0.52) | -8.6%<br>(-3.7% to -13.5%) |
| | Change in rate of incident disability post-stroke, relative to pre-stroke rate $\beta_3$ | 1.06<br>(0.93-1.20) | 1.00<br>(0.91-1.11) | -- |

AME represents the predicted absolute change based on the population’s comorbidities. AME for “change in rate of incident disability post stroke relative to pre-stroke rate” not estimated when the 95% CI for adjusted OR included 1.

## Appendix 5: Description of the method of recycled predictions to determine the contribution of strokes to population-level burden of disability

To estimate the contribution of strokes to the population disability burden, we determined the dependent-years had there been no strokes in the cohort.

First, we used the parameter estimates from the regression model (Appendix 4) to predict the probability of independence for each individual over 7.4 years given their specific covariates, if they had a stroke and when it occurred, and survey sampling weight. We did not project beyond 7.4 years because less than 25% of the cohort contributed more than 7.4 years of follow up time. For the 7.4% who had a stroke, this corresponds to the area of A + B. For the 92.6% that did not have a stroke, this corresponds to the area of A + B + C.

Next, among the 8.4% who had a stroke, we predicted the likelihood of independence assuming they did not have a stroke Area of A + B + C.

The difference between these two measures summed over the population represents the dependent-years attributable to stroke. We used this method to estimate the population burden for each of the three functional outcomes (ADL independence, IADL independence, community dwelling).

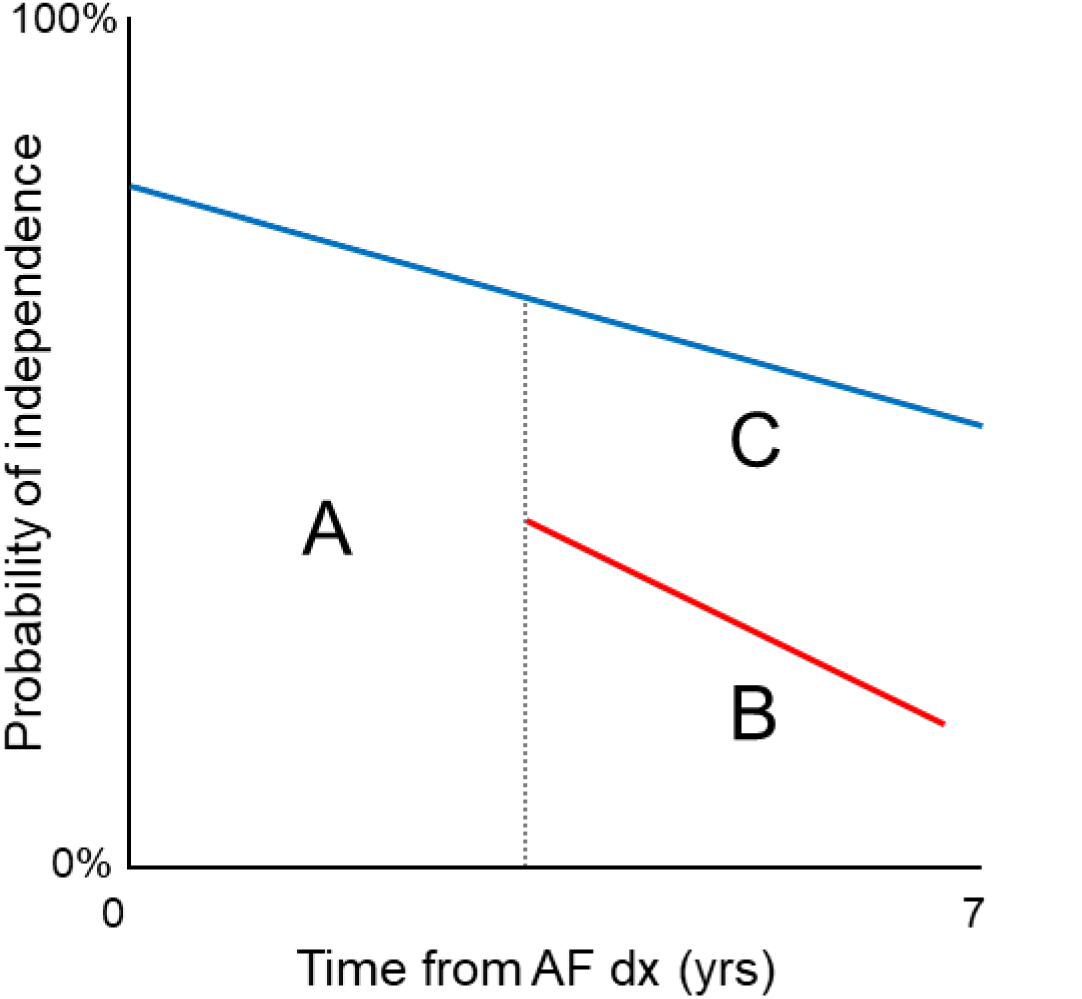

## Appendix 6: STROBE statement checklist

STROBE Statement—Checklist of items that should be included in reports of ***cohort studies***

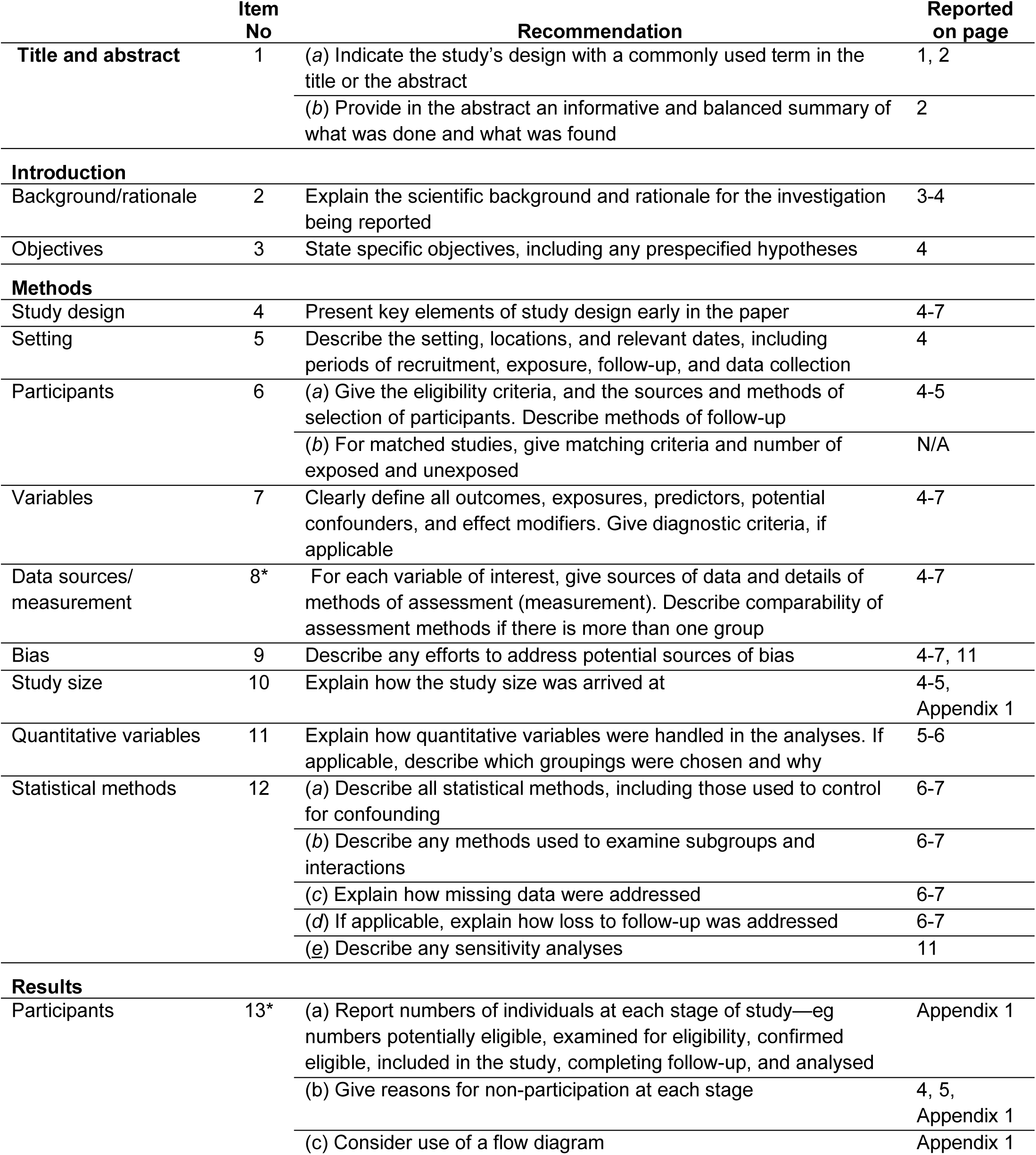
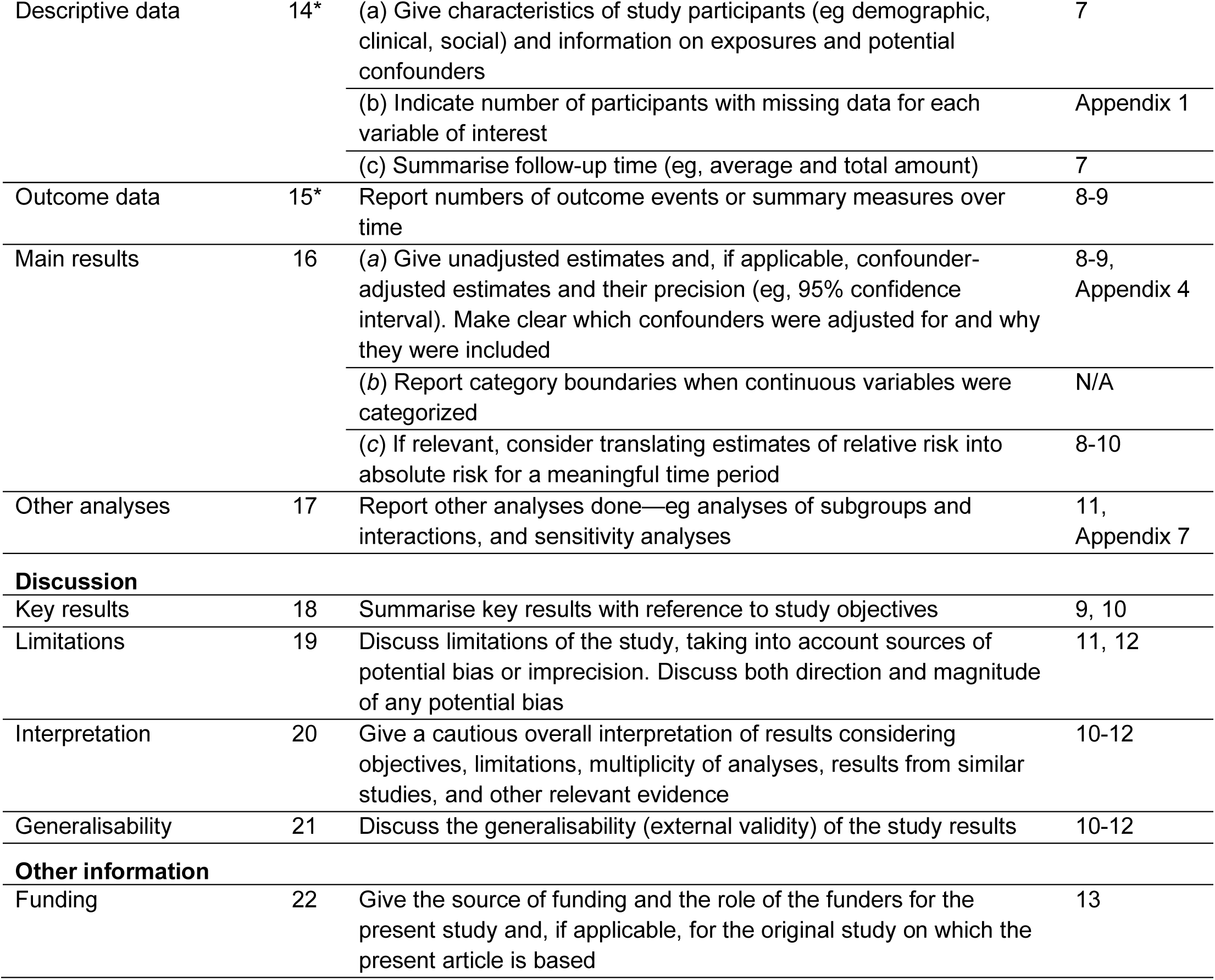

## Appendix 7: Rates of censoring by stroke status

*Participants were censored at the time of Medicare disenrollment, HRS drop out, or death, whichever came first. To address the possible bias that those with the most severe or deadly strokes would be more likely to be censored, we performed a sensitivity analysis assessing rates of censoring by year according to stroke status. As demonstrated above, there was no differential loss to follow-up by stroke status. Moreover, rates of censoring were similar between stroke versus non-stroke on a year-by-year basis.

| Censored/year* | No stroke (N=3268) |  | Stroke (N=262) |  |
| --- | --- | --- | --- | --- |
|  | N | % | N | % |
| 0-1 | 735 | 22% | 23 | 9% |
| 1-2 | 348 | 11% | 30 | 11% |
| 2-3 | 353 | 11% | 32 | 12% |
| 3-4 | 276 | 8% | 16 | 6% |
| 4-5 | 285 | 9% | 22 | 8% |
| 5-6 | 220 | 7% | 19 | 7% |
| 6-7 | 190 | 6% | 22 | 8% |
| 7-8 | 156 | 5% | 20 | 8% |
| 8-9 | 127 | 4% | 11 | 4% |
| 9-10 | 128 | 4% | 14 | 5% |
| 10-11 | 94 | 3% | 10 | 4% |
| 11-12 | 74 | 2% | 9 | 3% |
| 12-13 | 73 | 2% | 9 | 3% |
| 13-14 | 62 | 2% | 5 | 2% |
| 14-15 | 43 | 1% | 9 | 3% |
| 15-16 | 39 | 1% | 3 | 1% |
| 16-17 | 25 | 1% | 0 | 0% |
| 17-18 | 17 | 1% | 1 | 0% |
| 18-19 | 11 | 0% | 2 | 1% |
| 19-20 | 11 | 0% | 2 | 1% |
| >20 | 1 | 0% | 3 | 1% |
| Total | 3268 | 100% | 262 | 100% |

## Notes

### Author Declarations

Ethical approval: The Human Research Protection Program Institutional Review Board at the University of California, San Francisco, approved this study (IRB# 16-19185).

